# Spatial Decomposition of Longitudinal RNFL Maps Reveals Distinct Modes of Glaucomatous Progression with Structure–Function and Genetic Signatures

**DOI:** 10.64898/2026.04.09.26350387

**Authors:** Liyin Chen, Yan Zhao, Mousa Moradi, Mohammad Eslami, Mengyu Wang, Tobias Elze, Nazlee Zebardast

## Abstract

**Purpose:** To determine whether spatial decomposition of longitudinal retinal nerve fiber layer (RNFL) change maps reveals distinct modes of glaucomatous progression masked by conventional averaging, and to validate these modes through structure–function mapping and genetic association analysis.

**Methods:** Pixel-wise RNFL rates of change were computed from longitudinal optic disc OCT scans of 15,242 eyes (8,419 adults with primary open-angle glaucoma [POAG]; Massachusetts Eye and Ear, 1998–2023). A loss-only constraint zeroed all thickening values, reflecting the biological prior that adult RNFL does not regenerate. Non-negative matrix factorization decomposed these maps into spatial progression components (80% training set). Components were evaluated in a held-out set (20%) for retinotopic structure–function concordance, visual field (VF) progressor classification against global and quadrant RNFL rates, and enrichment of genetic association signals at established POAG loci.

**Results:** Six anatomically distinct progression patterns emerged, including diffuse circumferential loss, focal peripapillary defects, and arcuate bundle degeneration. Pattern-based models significantly outperformed global RNFL rate for classifying VF progressors (area under the curve, 0.750 [95% CI, 0.709–0.790] vs. 0.702; P = .0096) and explained additional variance in functional decline (Nagelkerke pseudo-R², 0.301 vs. 0.198; P = .0011). Structure–function mapping confirmed retinotopic coherence. Spatial phenotypes recovered stronger genetic signals than global rates at 85.3% of established POAG loci, suggesting they capture more biologically homogeneous endophenotypes of progression.

**Conclusions:** Glaucomatous structural progression occurs through spatially distinct modes with independent structure–function and genetic signatures that conventional RNFL averaging obscures.

## Introduction

Primary open-angle glaucoma (POAG) is structurally defined by localized loss of retinal nerve fiber bundles, resulting in characteristic spatial patterns of neurodegeneration rather than uniform, diffuse thinning. However, the prevailing clinical paradigm for monitoring structural progression paradoxically relies on averaging retinal nerve fiber layer (RNFL) thickness measurements globally or within fixed geometric quadrants. While convenient, this anatomical averaging effectively dilutes the signal of focal progression, obscuring localized neurodegenerative changes amidst the background noise of measurement variability and segmentation artifacts^1,2^. Consequently, this loss of spatial resolution not only reduces the sensitivity of clinical progression detection but also creates a phenotypic bottleneck that hinders the discovery of genetic factors underlying disease worsening.

To address the limitations of predetermined anatomical sectors, recent efforts have turned to data-driven approaches that utilize the entire spatial information of RNFL thickness maps. Non-negative Matrix Factorization (NMF) is a well-established unsupervised machine learning method that decomposes spatial data into additive, non-negative components that naturally yield “parts-based” representations^3^. Each component captures a distinct anatomical region, and their sum reconstructs the whole. NMF has been successfully applied to derive patterns of functional visual field loss and cross-sectional RNFL thickness archetypes^4,5^, revealing spatial patterns of damage that aligned with known nerve fiber bundle trajectories.

However, cross-sectional archetypes describe the cumulative state of structural damage at a single time point rather than the trajectory of active neurodegeneration. An eye exhibiting an inferotemporal damage archetype may currently be progressing superiorly, temporally, or diffusely. Characterizing progression thus requires decomposing not the static thickness maps, but their longitudinal change. A recent large-scale genome-wide association study (GWAS) of cross-sectional RNFL thickness identified only a single locus overlapped with established POAG susceptibility variants^6^. This disconnect highlights that global RNFL thickness at a single time point may be a noisy, nonspecific phenotype for identifying POAG genetic signals, reflecting heterogeneous baseline anatomy, measurement variability, and accumulated past injury. Instead, we hypothesize that rates and patterns of progressive thinning more directly capture the active neurodegenerative process and are therefore more likely to align with POAG-related genetic architecture.

In this study, we applied NMF to longitudinal RNFL rate-of-change maps to reveal distinct, biologically valid spatial phenotypes of POAG progression that were masked by conventional averaging. We enhanced the signal-to-noise ratio of the RNFL maps with a “loss-only” constraint to zero out thickening artifacts, which leveraged the biological prior that the adult RNFL does not regenerate.

Using a large longitudinal cohort of 15,242 eyes, we derived a spatial atlas of glaucomatous progression and validated its utility using 80/20 train-test split. We comprehensively assessed the clinical and biological validity of these spatial phenotypes through three distinct modalities: (1) anatomical coherence with retinotopic structure-function correlations; (2) prognostic utility for predicting functional visual field decline; and (3) genetic association analyses to determine if spatial phenotyping offers improved genetic tractability compared to global metrics.

## Methods

The Institutional Review Board at Massachusetts Eye and Ear Hospital approved this study with a waiver of consent to review retrospective data. The study adhered to the tenets of the Declaration of Helsinki and was compliant with the Health Insurance Portability and Accountability Act.

### Study Population

We considered adult patients followed at the Massachusetts Eye and Ear Hospital glaucoma clinic between 1998 and 2023 with a diagnosis of POAG as determined by International Classification of Diseases, Tenth Revision (ICD-10) code H40.1 and did not have a diagnosis code of secondary glaucoma, comorbid optic nerve or macular pathologies (Table S1). To maximize statistical power for the genetic association analyses, all patients with available genotype data were assigned to the validation cohort; the remaining patients were randomly partitioned to achieve the final 80:20 ratio.

Distinct inclusion criteria were applied to the training, VF, and genetic sub-cohorts to accommodate specific analysis requirements. The training sub-cohort required: (1) at least 3 high-quality optic nerve head (ONH) OCT scans; (2) age larger than 30 years old at OCT baseline; and (3) minimum 1-year follow-up. The validation cohort served as the parent group for two overlapping analyses: functional and genetic. The VF analysis sub-cohort required: (1) age larger than 30 years old at VF baseline; (2) at least 2 high-quality OCT scans and at least 5 reliable visual field (VF) tests; and (3) temporal concordance (scans within 1 year of VF). Both eyes were included in VF analyses if criteria were met. The genetic analysis sub-cohort was restricted to patients of predicted European descent (due to limited sample sizes in other ancestries, N < 100). For patients with bilateral data, the eye exhibiting greater cumulative progression (defined by the larger sum of pattern coefficients) was selected to enrich for phenotypic disease activity. Measurement bias was addressed through standardized quality control thresholds for OCT, VF, and genetic data.

### OCT Imaging and RNFL Thickness Map

All ONH OCT scans (Optic Disc Cube 200×200) were acquired using Cirrus HD-OCT (Carl Zeiss Meditec). Only scans with signal strength >6 were included. Automated quality control excluded scans with severe segmentation errors (fraction of pixels with negative thickness >0.90 or boundary coordinates at zero >0.85). Left eye images were mirrored to a right-eye orientation. To ensure accurate longitudinal pixel-to-pixel correspondence, RNFL thickness maps were registered by first registering their corresponding en-face fundus images, followed by applying the same transformation matrixes to the RNFL thickness maps. Registration was performed using SuperRetina^7^, a deep learning–based retinal feature extractor, with correspondences established via mutual nearest-neighbor matching and geometric transforms estimated using RANSAC. Registered images were further aligned to a common optic disc center (defined as the median centroid across all eyes) to ensure consistent spatial correspondence across the cohort. More registration details can be found in Supplementary Methods. To minimize edge artifacts and variations in disc size, we masked the central 1.8 mm (optic disc) and a 0.3 mm peripheral margin, yielding a 5.4 × 5.4 mm analysis region^4^. The thickness maps then underwent artifact correction as described in RNFLT2Vec^8^.

### Derivation of Spatial Progression Patterns

To isolate localized progression signals, we performed pixel-wise linear regression on longitudinal RNFL thickness maps. This yielded a slope map (β₁) representing the rate of change (μm/year) for every pixel. To focus exclusively on neurodegenerative thinning, a “loss-only” transformation was applied: β̂= max (0, - β₁). This transformation leverages a strong biological prior: in adult glaucoma, retinal ganglion cell axons do not regenerate, and therefore true RNFL thickening over time is not expected. Measured increases in pixel-wise thickness most commonly arise from segmentation variability, scan registration error, optical artifacts such as changes in signal penetration or co-occurring retinal and optic nerve pathology. By retaining only thinning rates, the loss-only constraint functions as a biologically informed denoising step that selectively suppresses the dominant non-glaucomatous source of longitudinal measurement variability^9,10^.

After generating the loss-only RNFL change maps for each eye, we combined the maps from the training set and applied NMF to discover latent spatial progression patterns. The optimal number of latent patterns (k) was selected from k = 2 to 20 based on convergent evidence from multiple criteria. Quantitatively, we identified an elbow in reconstruction error in the range between 4 to 10. Anatomically, k = 6 was the minimum rank that achieved separation of superior versus inferior hemiretinal damage and focal peripapillary versus distal arcuate bundle defects. Lower ranks (k < 6) conflated anatomically distinct defect patterns (e.g., merging inferior-temporal and inferior-arcuate loss), whereas higher ranks (k > 6) fractured spatially coherent bundles into redundant sub-components without improving prognostic performance for visual field progression (Supplementary Figure S2).

To obtain pattern coefficients for the validation set, each eye’s loss-only slope map was projected onto the fixed pattern matrix derived from the training set. This ensured that the pattern definitions were entirely independent of the validation data, preventing information leakage.

### Calculation of RNFL Thickness Change Slopes

We extracted global average RNFL thickness and quadrant thickness measurements (temporal, superior, nasal, inferior) from each OCT scan. Only scans with average RNFL thickness between 33 μm (3 standard deviation [SD] below the mean POAG eyes) and 126 μm (3 SD above the mean healthy eyes) were included, as values outside this range likely reflected imaging artifacts or segmentation errors. For eyes with multiple scans on the same date, the scan with the latest time stamp was selected. For each eye, ordinary least squares (OLS) linear regression was applied to calculate the rate of RNFL thinning. Time since baseline (in years) was used as the independent variable, and RNFL thickness (in micrometers) as the dependent variable. Separate regression models were fit for global average RNFL thickness and for each of the four quadrants.

### Visual Field Testing

VF tests in our study were obtained with a Humphrey Visual Field Analyzer (Carl Zeiss Meditec) using the Swedish Interactive Testing Algorithm Standard, Fast, or Faster strategies and the 24-2 test pattern. Only reliable VF tests were considered for analysis. VF tests with false-negative rates ≥ 20%, false-positive rates ≥20%, and fixation loss rates ≥33% were defined as unreliable. Eyes with at least 5 reliable VF tests were included for analyses.

Point-wise slopes (dB/year) were calculated by regressing the total deviation (TD) values at each location against time since baseline (average of first 2 tests) using ordinary least squares linear regression, yielding a slope representing the rate of sensitivity change at each test location in the 24-2 grid. The two blind spot locations (points 26 and 35) were excluded, resulting in 52 location-specific slopes per eye.

We additionally assessed visual field progression using four complementary VF progression metrics, namely, MD slopes, VFI slopes, Collaborative Initial Glaucoma Treatment Study (CIGTS)^11^ and Advanced Glaucoma Intervention Study (AGIS)^12^ algorithms. To maximize specificity and establish a robust ground truth for validation, we defined consensus progressors as eyes meeting at least three of these four criteria.

The MD slope (dB/year) and the Visual Field Index (VFI) slope of each eye were calculated by regressing the MD values and the VFI values, respectively, against time since baseline using OLS linear regression. MD progression was defined with MD slopes less than -0.5 dB/year and the P value was less than 0.05. VFI progression was defined with VFI slopes less than -1.0 %/year and the P value was less than 0.05. In addition to trend-based analyses, we assessed VF progression using two established event-based scoring systems. The AGIS score (range 0–20) quantifies VF defect severity based on the number, depth, and spatial clustering of depressed test locations, with separate components for the superior hemifield (0–9 points), inferior hemifield (0–9 points), nasal step (0–1 point), and central involvement (0–1 point). Clusters were defined as 3 or more adjacent points with TD values ≤ −5 dB, with at least one point reaching moderate depression (TD ≤ −10 dB). AGIS progression was defined as an increase of ≥4 points from baseline confirmed on 3 consecutive visits. The CIGTS score (range 0–20) was calculated by transforming the total deviation probability values at each test location into ordinal scores (0–4 points based on probability thresholds of 0.05, 0.02, 0.01, and 0.005), summing across all 52 non-blind-spot locations, and normalizing to a 0–20 scale. CIGTS progression was defined as an increase of ≥3 points from baseline confirmed on 2 consecutive visits. For both methods, baseline scores were calculated as the mean of the first 2 visits.

## Genotype data

A subset of 842 MEE patients had available genotype data for performing genetic analyses. This genotype data was part of the Mass General Brigham Biobank (MGBB)^13^. Samples were genotyped in four batches using biobank SNP arrays offered by Illumina. Standard quality control procedures were applied at both sample and variant levels. Details can be found in Supplementary Methods.

## Statistical Analysis

To assess anatomical validity, we calculated Spearman rank correlations between pattern weights and spatially corresponding metrics: sectoral RNFL thinning rates and point-wise VF total deviation slopes. For all analyses involving pattern weights and VF metrics, we inverted the sign of pattern weights so that negative values indicated faster thinning, aligning with clinical convention for RNFL and VF change rates.

To evaluate whether patterns independently stratified functional decline, we compared MD slopes between eyes with high (top 20th percentile) versus low (bottom 20th percentile) pattern loadings for each of the six patterns. Multivariable linear regression adjusted for age, sex, and race was used to estimate mean differences, with robust cluster-adjusted standard errors accounting for bilateral eye inclusion. P values were corrected using the Benjamini-Hochberg false discovery rate.

We evaluated discrimination for classifying VF progressors using nested logistic regression models with L2 regularization: (1) covariates only (age, sex, race); (2) covariates plus global RNFL rate; (3) covariates plus quadrant RNFL rates; (4) covariates plus pattern weights; (5) covariates plus pattern weights and global RNFL rate; and (6) covariates plus pattern weights and quadrant RNFL rates. Models were fit using balanced class weights with patient-clustered 5-fold cross-validation to prevent data leakage from bilateral eyes. Discrimination was quantified by area under the receiver operating characteristic curve (AUC), integrated discrimination improvement (IDI)^14^, and Nagelkerke pseudo-R². Confidence intervals for AUC were derived via patient-level bootstrap resampling (2000 iterations); model comparisons used permutation testing (2000 permutations).

Genome-wide association analyses were performed for each pattern weight and for global RNFL thinning rate among 645 participants of predicted European ancestry using REGENIE (cite). A Bonferroni-adjusted significance threshold of P < 8.3 × 10⁻⁹ (5 × 10⁻⁸ / 6 patterns) was applied for pattern GWAS, whereas the standard P < 5 × 10⁻⁸ was applied for global rate. Linkage disequilibrium–independent loci were identified by clumping (500-kb window, r² < 0.1) using the European 1000 Genomes reference panel. One hundred and nine established POAG risk variants from multi-ancestry GWAS meta-analyses^15^ that passed quality control in our data were used to compare the maximum |Z|-score across patterns versus global rate at each locus to assess signal enhancement independent of threshold selection.

All hypothesis tests were two-sided. Samples with missing covariate data were excluded.

## Statistical Software

Image analyses and statistical comparisons were implemented in Python 3 using NumPy (numerical operations), pandas (data manipulation), SciPy (statistical tests and linear regression via scipy.stats.linregress), scikit-learn (NMF implementation and machine learning models), PyTorch (image preprocessing), and matplotlib/seaborn (visualization). Genetic analyses were performed using REGENIE^16^ for association testing and PLINK v2.0^17^ for clumping.

## Results

### Study population

From an initial database of 21,179 adult POAG eyes (10,807 patients) with longitudinal OCT measurements, 15,242 eyes (8,419 patients) passed strict image quality control and registration requirements. This eligible pool was split at the patient level into training (80%) and validation (20%) cohorts. Within the training cohort, 8,705 eyes met the stricter longitudinal inclusion criteria (having at least 3 scans over a minimum of 1 year) and were used for pattern derivation.

The validation cohort was used for independent structure-function and genetic assessments. Of the eyes assigned to validation, 1,232 eyes (789 patients) met the requirements for VF analysis, and 1,704 eyes (842 patients) had quality-controlled genotype data. Table 1 summarizes the demographic and clinical characteristics of the study population. Importantly, markers of disease activity, including baseline global RNFL thickness, baseline VF MD, global RNFL thinning rate, and MD slope, did not differ statistically between the training and validation cohorts (P > 0.05 for all), confirming that the partition produced representative datasets.

**Table 1.**
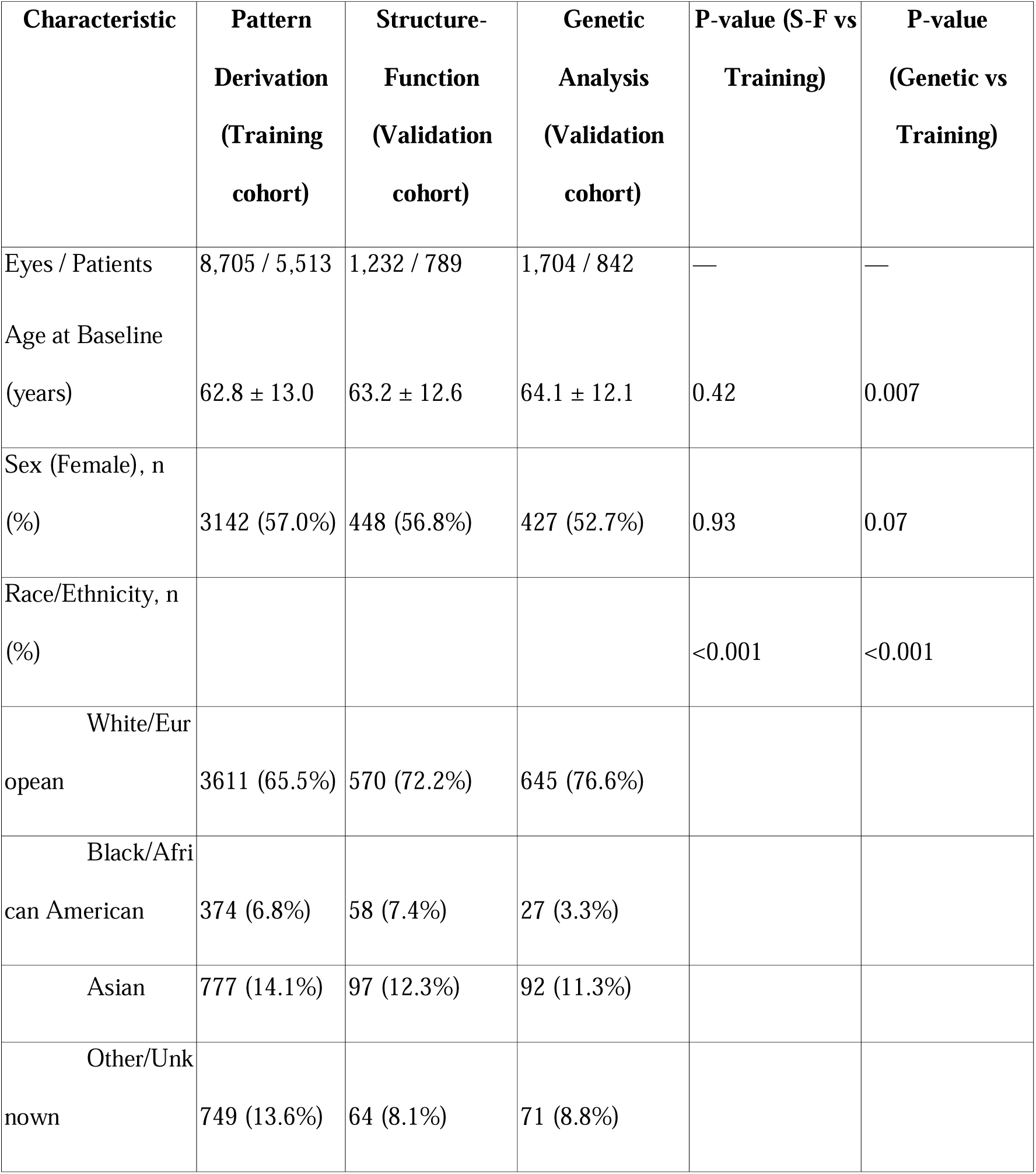
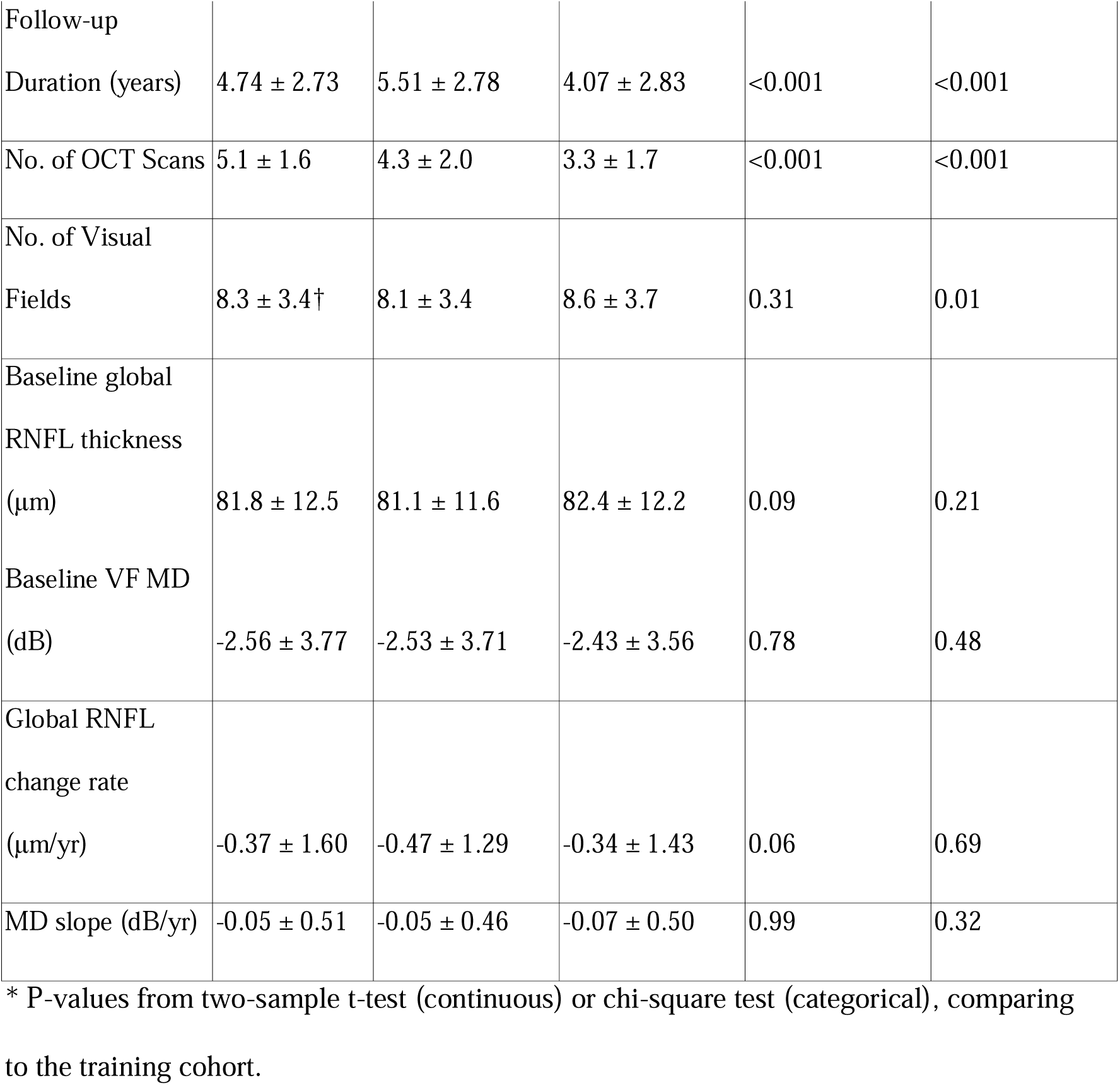
Demographic and clinical characteristics of the study population.

### POAG progression patterns

Figure 1 illustrates the pipeline for deriving spatial progression phenotypes. For each eye, pixel-wise linear regression of longitudinal RNFL thickness maps yielded a rate-of-change map quantifying local thinning or thickening over time. A loss-only transformation then set all thickness-increasing pixels to zero, isolating true neurodegenerative loss from measurement noise and segmentation artifacts. NMF was applied to the combined loss-only maps from all training eyes, decomposing them into six spatial basis patterns that collectively form an atlas of glaucomatous progression. Each basis pattern (Figure 1, bottom) represents a spatial template of RNFL loss, where brighter regions indicate areas that contribute more strongly to that pattern. Any individual eye’s progression can be expressed as a weighted sum of these six patterns, with higher weights indicating greater loading of that spatial phenotype. The pattern weights thus can be served as quantitative endophenotypes characterizing the spatial configuration of each eye’s neurodegeneration.

**Figure 1.**
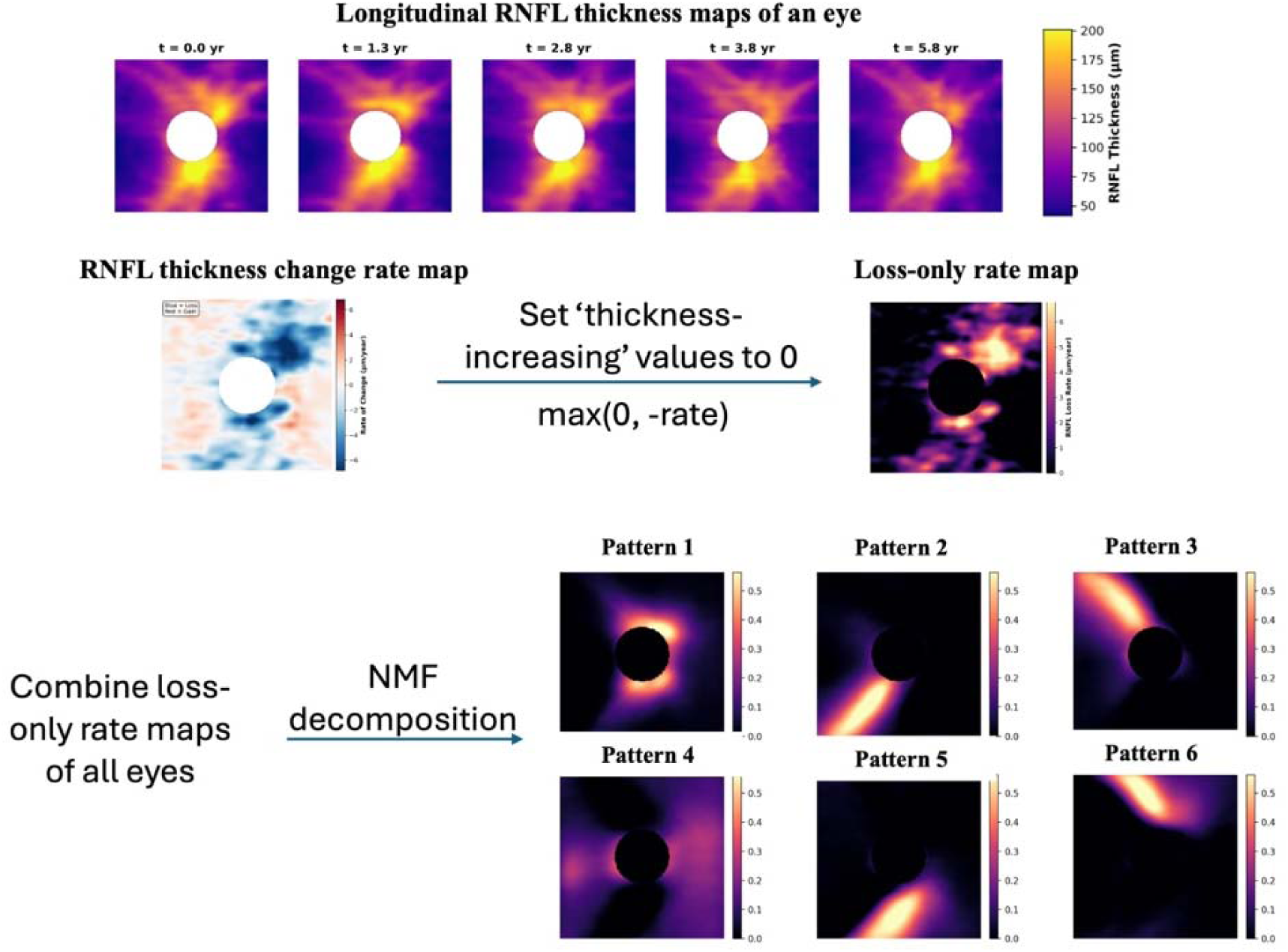
Overview of the pipeline for identifying latent spatial patterns of glaucomatous progression from longitudinal OCT data. Top: Representative longitudinal retinal nerve fiber layer (RNFL) thickness maps from a single eye over 5.8 years of follow-up, demonstrating progressive thinning in the superior and inferior arcuate regions. All maps were registered to ensure pixel-wise spatial correspondence across timepoints. The central white circle denotes the masked optic disc region. Color scale: RNFL thickness in micrometers (μm). Middle: For each eye, pixel-wise ordinary least squares regression yielded a rate-of-change map (left; blue = thinning, red = thickening). A “loss-only” transformation set thickness-increasing pixels to zero (right), isolating neurodegenerative thinning from measurement noise and segmentation artifacts. Bottom: Non-negative matrix factorization (NMF) decomposed the combined loss-only maps from all training eyes into six spatially distinct, additive basis patterns. These learned patterns correspond to recognizable anatomical structures: diffuse circumferential loss (Pattern 1), focal inferior-temporal and superior-temporal peripapillary loss (Patterns 2 and 3), a low-intensity background/noise component (Pattern 4), and inferior and superior arcuate bundle degeneration extending into the temporal periphery (Patterns 5 and 6). Each eye’s progression can be represented as a weighted combination of these six spatial phenotypes.

Six spatially distinct patterns emerged. Pattern 1 demonstrated diffuse circumferential RNFL loss surrounding the ONH, with involvement of both superior and inferior regions. Patterns 2 and 5 both involve the inferior hemiretina but differ spatially. Specifically, pattern 2 represented focal peripapillary inferior-temporal thinning, while Pattern 5 followed the arcuate nerve fiber bundle trajectory extending into the temporal periphery. Similarly, Pattern 3 showed focal superior peripapillary loss whereas Pattern 6 indicated superior arcuate bundle-following degeneration. Pattern 4 showed the lowest intensity of RNFL change across in a diffuse manner, likely represents a background component capturing residual measurement variability.

## Clinical validation

### Spatial Concordance with Structural and Functional Progression

To confirm that the derived patterns exhibit anatomically coherent structure-function relationships in independent data, we projected the pattern basis onto the held-out validation set and computed pattern coefficients for each eye. We then examined correlations between these coefficients and both sector-specific RNFL progression rates and point-wise VF sensitivity slopes.

Pattern weights correlated most strongly with RNFL progression in the expected anatomical sectors (Supplementary Figure S2). Specifically, patterns 2 and 5 (inferior-temporal RNFL damage) show the largest correlation with inferior sector RNFL progression than other sectors, followed by with the temporal sector. Similarly, pattern 3 and 6 showed strong correlations with the superior sector progression rates. Pattern 1 (diffuse circumferential loss) correlated broadly across multiple sectors, while Pattern 4 (minimal change) demonstrated a moderate correlation specifically with the nasal RNFL sector.

Structure-function analysis also revealed spatially specific relationships consistent with retinotopic anatomy (Figure 2). Patterns representing inferior RNFL damage (Patterns 2 and 5) correlated preferentially with the pointwise TD change rates in the superior hemifield, while Pattern 3 and 6 (superior RNFL damage) preferentially correlated with inferior hemifield decline. This reciprocal hemifield correspondence reflects the known projection of retinal nerve fibers across the horizontal raphe^18^. While Patterns 2 and 5, and similarly Patterns 3 and 6, affected the same hemiretina, Pattern 2 and 3 represented paracentral damage with likely involvement of the papillomacular axons, whereas Pattern 5 and 6 captured arcuate bundle defects. Pattern 4 showed negligible correlation with functional decline across the visual field. While our patterns demonstrate strong retinotopic coherence, the correlation is not absolute. This is consistent with the known non-linear relationship between structural loss and functional decay^19^, as well as the temporal lag often observed between structural thinning and detectable perimetric change^20^.

**Figure 2.**
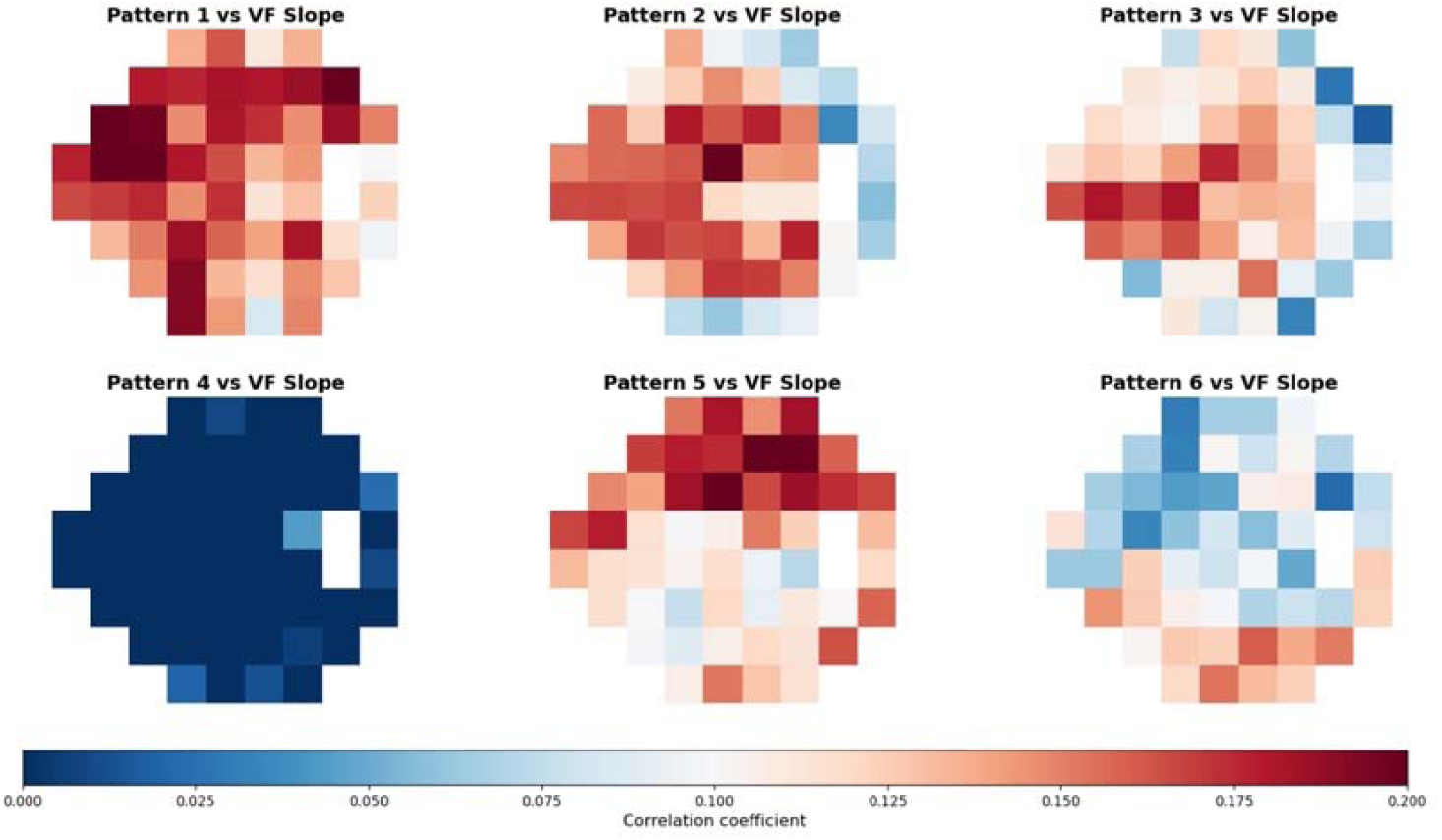
Heatmaps displaying the spatial correlation between pattern coefficients and point-wise VF TD slopes in the independent validation set. Red color indicates greater correlation.

### Pattern-Specific Associations with Functional Decline

To examine the relationship between structural patterns and functional loss, we compared the mean MD progression rates between eyes with large pattern coefficients (top 20%) versus smaller pattern coefficients (bottom 20%) for each pattern. Five of the six structural patterns were significantly associated with accelerated functional decline (Figure 3) (Supplementary Table S2). Pattern 1 (diffuse peripapillary loss) demonstrated the strongest discrimination. Specifically, eyes with high coefficients of Pattern 1 exhibited a mean MD slope of −0.13 dB/year, whereas those with low expression remained functionally stable (+0.02 dB/year), resulting in a significant adjusted mean difference of −0.15 dB/year. Pattern 5 (inferior arcuate) showed the second strongest association, with high-expression eyes progressing significantly faster than low-expression eyes (mean difference: −0.081 dB/year). Similarly, high expression of Patterns 2 (inferotemporal), 6 (superior arcuate), and 3 (superotemporal) was associated with significantly faster functional decay, with mean differences ranging from −0.038 to −0.062 dB/year, all of which passed statistical significance of FDR-adjusted P < 0.05 (except for Pattern 3, which pass FDR-adjusted P < 0.1). These pattern-based stratification signal was robust even against the background of high variability in MD slopes (Cohort mean: -0.05 ± 0.46 dB/year). Pattern 4 was the only phenotype that did not significantly stratify functional progression. suggesting that this component likely represents a baseline or residual signal.

**Figure 3.**
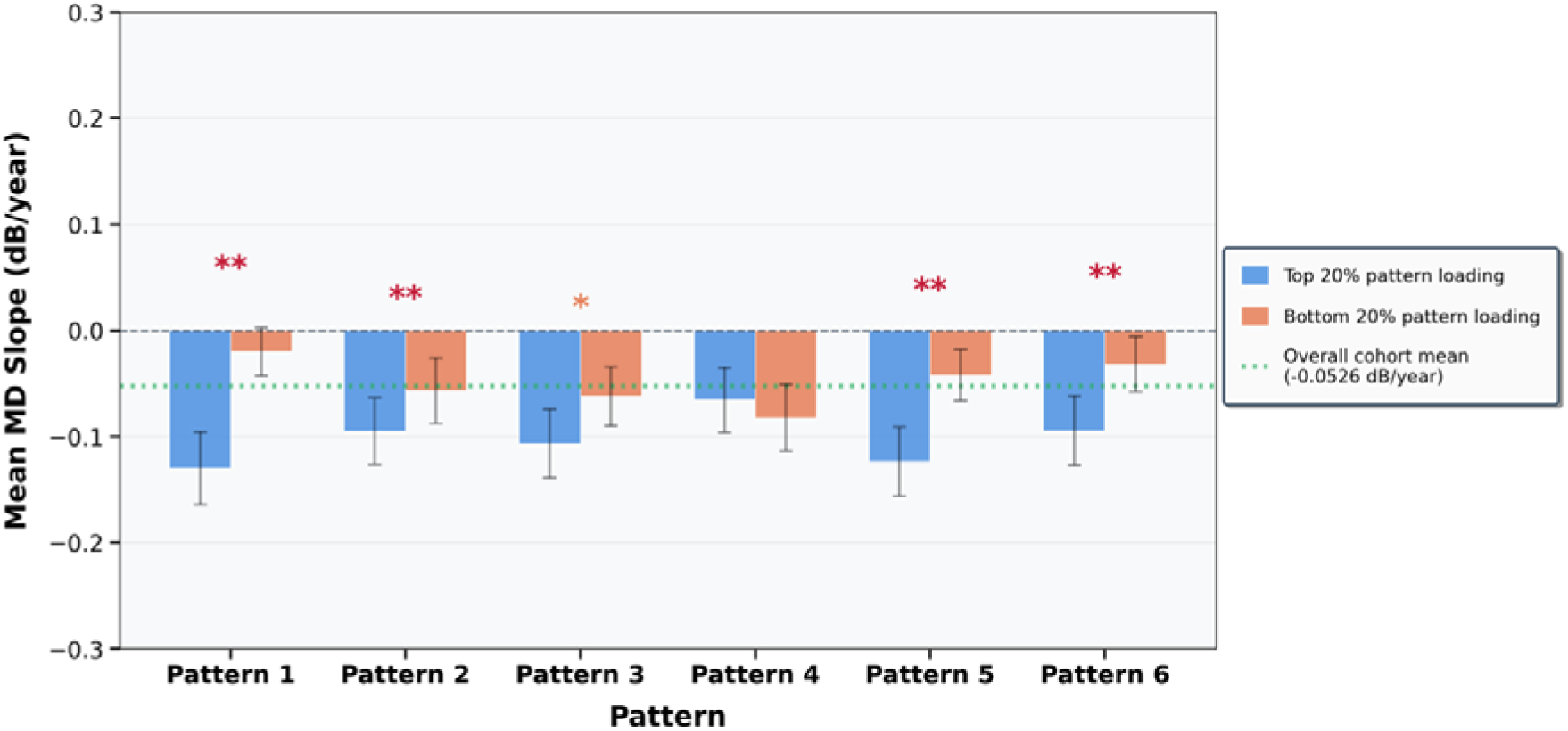
Pattern-based risk stratification of mean MD slope. Comparison of mean MD slopes (dB/year) between eyes with high (top 20th percentile, blue) versus low (bottom 20th percentile, red) pattern coefficients for each spatial pattern. Error bars represent 95% confidence intervals derived from patient-level bootstrapping. The dotted green line represents the overall cohort mean MD slope. The ‘*’ annotation indicates statistically significant differences in the mean MD slopes the two groups for each pattern after adjustment for age, sex, and race. ** indicates FDR-adjusted P < 0.05; * indicates FDR-adjusted P < 0.10.

### Performance of NMF patterns in VF progressor classification

In the same held-out validation set, using patterns as predictors for POAG progression (defined as a consensus of four VF progression metrics described in Methods) led to better classification performance than anatomical averages and provided additive prognostic value when combined with conventional metrics. When evaluating the ability of conventional metrics and the progression patterns to classify POAG progressors and non-progressors, using the patterns as predictors led to a 6.8% and 4.0% increase in AUC compared to using global RNFL rate or sectoral RNFL change rates as predictors, respectively (Figure 4a) (Supplementary Table S3). The pattern-based model also explained 10.2% more variance in functional progression (pseudo-R^2^ = 0.301 vs 0.199 for global rate) (Figure 4b) and yielded an IDI of 0.071 relative to the global model (P < 0.005 for both). Furthermore, NMF patterns provided distinct, additive predictive power when combined with conventional measurements. Models that combined patterns with global or sectoral RNFL rates, explained an additional 10.4% and 12.6%, respectively of the variance in functional progression as compared to global RNFL slopes.

**Figure 4.**
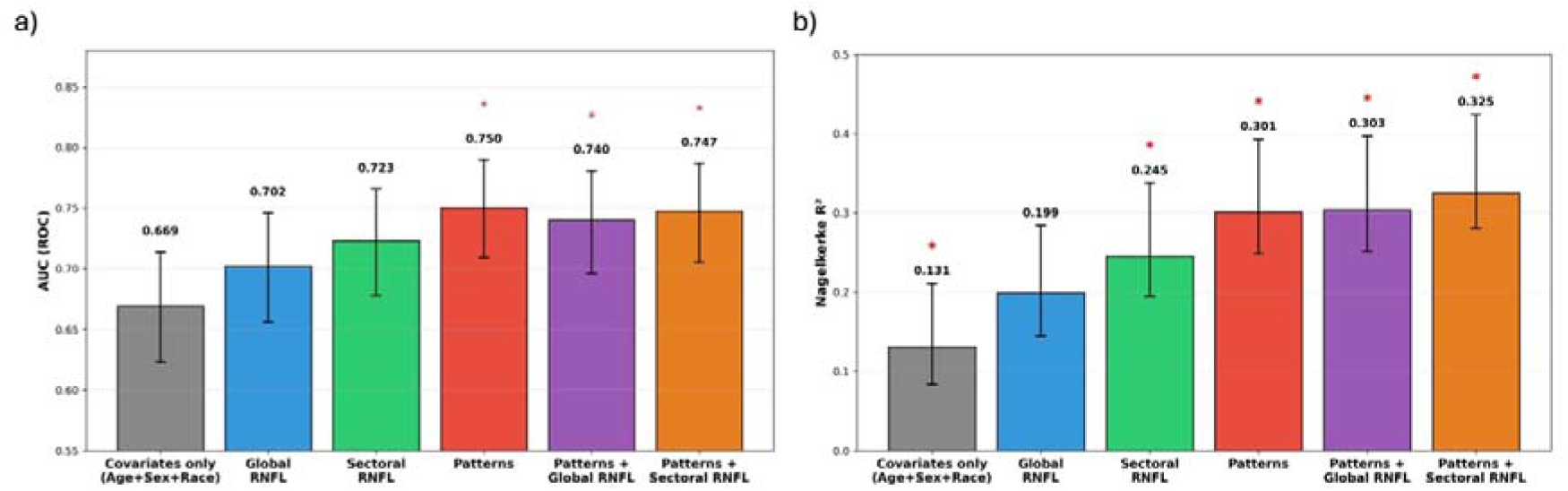
Prognostic Performance for Visual Field Progression in the Validation Set. Bar charts comparing the discrimination and goodness-of-fit for logistic regression models classifying eyes as VF progressors vs. non-progressors. A) Area Under the Receiver Operating Characteristic Curve (AUC). B) Nagelkerke R^2^ values indicating the proportion of variance explained by each model. The model using NMF patterns alone (Red) significantly outperforms models using Global (Blue) or Sectoral (Green) RNFL thinning rates. The combination of Patterns with Global RNFL rate (Purple) further improved discrimination. The comprehensive model combining NMF patterns with Sectoral rates (Orange) achieved the highest predictive power. * Indicates a statistically significant improvement P < 0.005) compared to the global RNFL rate model. Error bars indicate 95% confidence intervals.

### Biological validation with genetic data

To evaluate whether the NMF-derived spatial endophenotypes offer enhanced genetic tractability compared to conventional metrics, we performed genome-wide association studies for each of the six progression patterns and for global RNFL thinning rate in participants with available genotype data (645 European descent samples and 7,804,018 variants after quality control).

Despite the modest sample size, progression patterns yielded substantially more genome-wide significant associations than global RNFL rate. There were 23 LD-independent loci associated with at least one NMF pattern (P < 5 × 10^-8^), where 9 of them passed the more stringent Bonferroni-adjusted threshold accounting for the 6 patterns (P < 8.3 × 10⁻⁹). In contrast, only 4 variants reached genome-wide significance (P < 5 × 10⁻⁸) for global RNFL thinning rate. At the suggestive significance level (P < 1 × 10⁻^6^), this disparity was similarly pronounced, where 111 LD-independent variants were associated with progression patterns (40 after Bonferroni correction for the 6 patterns), compared with 23 variants for global rate. These findings indicate that spatial decomposition isolates progression signals with stronger genetic associations than anatomically averaged metrics, consistent with the hypothesis that NMF patterns represent more biologically homogeneous and therefore more genetically tractable phenotypes. There was no evidence of systematic inflation, with genomic inflation factors (λ) close to unity across all GWAS (mean λ = 1.00; range: 0.98–1.01; Supplementary Figure S3).

To further validate the genetic relevance of the patterns, we examined associations at 109 established POAG susceptibility loci. Given the absence of large-scale GWAS specifically for glaucoma progression, established susceptibility loci provide the most robust available biological ground truth. We hypothesized that valid progression phenotypes should share genetic architecture with disease susceptibility; therefore, a more refined progression phenotype should recover these susceptibility signals more effectively than noisy averages. Across tested loci, the maximum association magnitude (|Z|) across the spatial patterns exceeded that of the global RNFL rate in 93 of the 109 (85.3%) tested variants. This finding demonstrated that although these loci were discovered for disease susceptibility, their effects on structural degeneration were more effectively captured by spatial progression phenotypes than by global RNFL rates.

### Illustrative case: Detection of focal progression masked by global RNFL stability

To illustrate the clinical utility of spatial phenotyping, we present a case from the validation cohort in which conventional global RNFL metrics failed to detect progression that was captured by our pattern analysis (Figure 6). This patient was followed for longer than 5 years with serial OCT and visual field testing. Despite exhibiting clear functional deterioration (MD declining from −9.5 to −16.2 dB; slope: −1.26 dB/year, P < 0.001), the global average RNFL thickness remained stable throughout follow-up (range: 68.3–72.8 μm; slope: +0.42 μm/year), and this eye would not have been identified as a structural progressor using conventional criteria.

**Figure 5.**
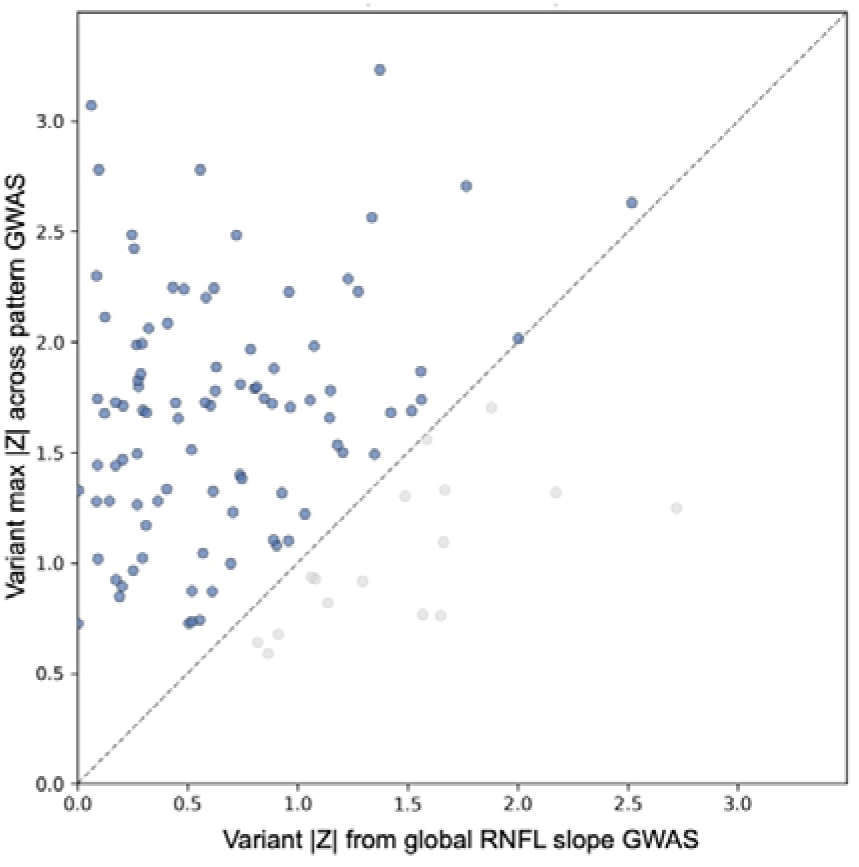
Comparing the association strength (Z-score) of the global RNFL rate (x-axis) versus the NMF pattern yielding the maximum absolute association (y-axis) for each established POAG risk variants. Points above the diagonal line indicate instances where isolating the spatial phenotype recovered a stronger genetic signal than anatomical averaging. Pattern-based GWAS recovered high association strength than global RNFL slope GWAS for 93 out of 109 known POAG loci.

**Figure 6.**
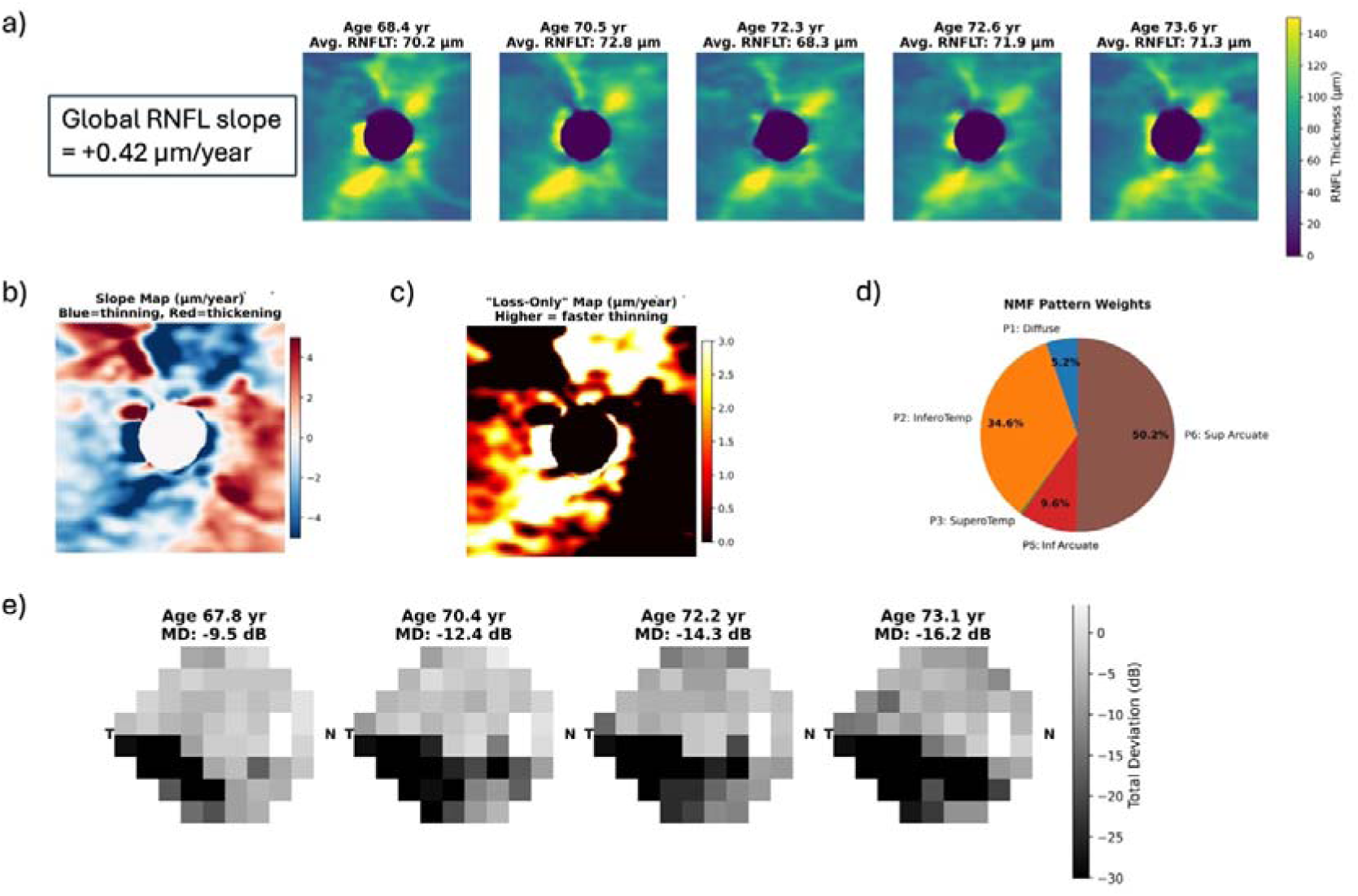
Case example demonstrating detection of focal progression masked by global RNFL stability. (A) Longitudinal RNFL thickness maps from a patient their 60’s with primary open-angle glaucoma followed over 5 years. Global average RNFL thickness fluctuated between 68.3 and 72.8 μm with an overall positive slope (+0.42 μm/year). (B) Pixel-wise slope map reveals focal superior-temporal thinning (blue) offset by regions of apparent thickening (red), likely representing segmentation or registration artifacts. (C) Pixel-wise slope map after application of the loss-only transformation. (D) NMF of the loss-only map identifies Pattern 6 (Superior Arcuate) as the dominant progression phenotype, with secondary contribution from Pattern 2 (Inferotemporal). (E) Corresponding visual field total deviation maps demonstrate progressive functional decline (mean deviation: −9.5 to −16.2 dB; slope: −1.26 dB/year, P < 0.001)

Examination of the pixel-wise slope map (Figure 6b) reveals why global averaging failed: regions of superior-temporal thinning (blue) were arithmetically offset by regions of apparent thickening (red), likely representing segmentation or registration artifacts. Application of the loss-only transformation (Figure 6c) isolated the neurodegenerative signal by removing non-biological thickening. NMF decomposition of this loss-only map (Figure 6d) identified Pattern 6 (Superior Arcuate) as the dominant progression phenotype, accounting for 50.2% of the total pattern weight, with secondary contribution from Pattern 2 (Inferotemporal, 34.6%).

Critically, the spatial pattern of structural loss corresponded precisely to the functional deficit: the patient’s visual field maps (Figure 6e) demonstrate progressive inferior hemifield loss, consistent with damage to the superior arcuate nerve fiber bundle captured by Pattern 6. This case exemplifies how spatial phenotyping can identify clinically meaningful progression in eyes that appear stable by global RNFL metrics.

## Discussion

In this study, we identified six spatially distinct phenotypes of glaucomatous RNFL progression through unsupervised decomposition of longitudinal thickness maps and validated their clinical and biological relevance in independent data. The central clinical finding is that the spatial configuration of structural loss carries prognostic information that is invisible to conventional global and sectoral averaging. In our validation cohort, pattern-based classification improved the identification of visual field progressors by 6.8% in AUC over global RNFL metrics. Our case example in Figure 6 illustrated how a patient with clear functional deterioration (MD declining from −9.5 to −16.2 dB) could appear structurally stable when assessed by global RNFL thickness alone, while spatial pattern analysis correctly identified the dominant superior arcuate progression phenotype. These results argue that clinical progression monitoring should incorporate spatial information beyond simple thickness averaging. By identifying the subset of patients whose focal progression is arithmetically masked by global averaging, spatial phenotyping may flag patients at risk of delayed treatment intensification who would otherwise be falsely reassured by stable summary metrics.

The derived patterns align with known neuroanatomy. Patterns representing inferior RNFL damage (Patterns 2 and 5) correlated preferentially with superior hemifield functional decline, and vice versa, consistent with the retinotopic projection of retinal nerve fibers across the horizontal raphe^18^. The inferior-temporal phenotypes spatially correspond to the macular vulnerability zone described by Hood et al.^21^, and the distinction between peripapillary focal loss (Patterns 2, 3) and distal arcuate bundle degeneration (Patterns 5, 6) recapitulates the clinical observation that early glaucomatous damage can present as either a focal wedge defect or an arcuate scotoma extending peripherally^1^. The recovery of these established anatomical relationships from an entirely data-driven, unsupervised analysis provides confidence that the patterns reflect genuine biological structure rather than statistical artifacts.

A major challenge in glaucoma progression monitoring and genetic discovery is the substantial measurement variability inherent in RNFL thickness and MD measurements, where noise often obscures the true neurodegenerative signal. Despite significant advances in mapping POAG susceptibility loci^22,23^, the genetic architecture of POAG progression remains largely elusive, a gap attributable in part to the substantial phenotypic noise inherent in conventional metrics^24^. Our work addressed this by incorporating a “loss-only” constraint to effectively separate pathological thinning from measurement artifacts. This constraint is grounded in the established understanding that retinal ganglion cell axon loss in glaucoma is irreversible; the adult mammalian RNFL does not regenerate^9,25^. Measured increases in pixel-wise thickness over time most commonly reflect segmentation variability, registration error, and optical artifacts rather than true biological thickening^10,26^. By filtering out non-biological noise, our method derived coherent spatial phenotypes that showed strong performance in prognostic classification and genetic tractability.

Our preliminary genetic analysis served as biological validation rather than as a GWAS discovery effort and should be interpreted accordingly given our modest sample size (645 European-ancestry participants). Nonetheless, the finding that spatial phenotypes yielded stronger association signals than global RNFL rate at 85.3% of established POAG susceptibility loci is noteworthy. This result is consistent with our hypothesis that spatial patterns represent more biologically homogeneous endophenotypes that are closer to the underlying genetic architecture than anatomically averaged metrics^27^. The disparity in genome-wide significant loci (23 across patterns vs. 4 for global rate) further supported this interpretation, though we emphasized that these associations require replication in larger cohorts before any locus-specific claims can be made. We envision that application of this framework to multi-center cohorts with dense genotyping will enable discovery of progression-specific genetic signals that remain hidden by the phenotypic noise of conventional averaging, ultimately facilitating genotype-guided risk stratification for POAG progression

The computational requirements for clinical implementation are minimal. Once the six-pattern basis matrix is established, scoring a new patient requires only a non-negative least squares projection, a calculation that executes in milliseconds. This efficiency may facilitate integration into clinical OCT software platforms, particularly as these systems increasingly incorporate computational analysis pipelines.

This study has several limitations that should be considered. First, while we utilized a rigorous 80/20 train-test split to ensure robustness, the analysis was conducted at a single tertiary center using one OCT platform. The generalizability of both the derived patterns and the performance gains to other clinical settings, patient populations, and OCT devices remains to be established through external validation. Second, the pixel-wise linear regression assumes a constant rate of RNFL change over the observation period; eyes with accelerating, decelerating, or episodic progression may be poorly characterized by this model, and future work should explore nonlinear or change-point approaches. Third, the genetic analyses were limited to participants of European ancestry due to sample size constraints, precluding assessment of cross-ancestry generalizability and limiting the applicability of our findings to diverse populations. With the continued multicenter collaborative genotyping efforts, we envision expanded genetic studies with diverse populations soon, which could enable discovery of POAG progression genetics and genotype-guided risk stratification.

In conclusion, unsupervised spatial phenotyping of longitudinal RNFL thickness maps reveal distinct modes of glaucomatous progression that are obscured by conventional averaging. These phenotypes demonstrate anatomical validity, improve prediction of functional decline, and show enhanced genetic tractability, supporting a precision medicine framework for glaucoma monitoring. The findings argue for a shift in how structural progression is conceptualized clinically: from “how fast is this eye thinning?” to “where and in what spatial configuration is neurodegeneration occurring?” The derived atlas provides a framework for characterizing progression heterogeneity that may ultimately inform personalized monitoring strategies and illuminate the biological pathways underlying different modes of glaucomatous neurodegeneration.

## Data availability statement

The pattern atlas matrix and NMF training code will be deposited in a public repository upon publication. Due to privacy restrictions and institutional data use agreements, individual-level clinical and genetic data cannot be shared publicly but may be available from the corresponding author upon reasonable request, subject to institutional review board approval and data use agreement.

## Acknowledgments

We are grateful to the patients of the MEE glaucoma clinic whose contributions were essential for this research.

## Funding

National Institutes of Health grants R01 EY036518 (NZ)

## Notes

### Competing Interest Statement

The authors have declared no competing interest.

## References

1. Lee EJ, Kim TW, Weinreb RN, Park KH, Kim SH, Kim DM. Trend-based analysis of retinal nerve fiber layer thickness measured by optical coherence tomography in eyes with localized nerve fiber layer defects. Invest Ophthalmol Vis Sci. 2011;52(2):1138–1144. doi:10.1167/iovs.10-5975

2. Leung CK shun, Cheung CYL, Weinreb RN, et al. Evaluation of retinal nerve fiber layer progression in glaucoma: a study on optical coherence tomography guided progression analysis. Invest Ophthalmol Vis Sci. 2010;51(1):217–222. doi:10.1167/iovs.09-3468

3. Lee DD, Seung HS. Learning the parts of objects by non-negative matrix factorization. Nature. 1999;401(6755):788–791. doi:10.1038/44565

4. Wang M, Shen LQ, Pasquale LR, et al. An Artificial Intelligence Approach to Assess Spatial Patterns of Retinal Nerve Fiber Layer Thickness Maps in Glaucoma. Transl Vis Sci Technol. 2020;9(9):41. doi:10.1167/tvst.9.9.41

5. Elze T, Pasquale LR, Shen LQ, Chen TC, Wiggs JL, Bex PJ. Patterns of functional vision loss in glaucoma determined with archetypal analysis. J R Soc Interface. 2015;12(103):20141118. doi:10.1098/rsif.2014.1118

6. Currant H, Hysi P, Fitzgerald TW, et al. Genetic variation affects morphological retinal phenotypes extracted from UK Biobank optical coherence tomography images. PLoS Genet. 2021;17(5):e1009497. doi:10.1371/journal.pgen.1009497

7. Jiazhen Liu, Xirong Li, Qijie Wei, Jie Xu, Dayong Ding. Semi-Supervised Keypoint Detector and Descriptor for Retinal Image Matching. https://arxiv.org/pdf/2207.07932

8. Shi M, Tian Y, Luo Y, Elze T, Wang M. RNFLT2Vec: Artifact-corrected representation learning for retinal nerve fiber layer thickness maps. Med Image Anal. 2024;94:103110. doi:10.1016/j.media.2024.103110

9. Weinreb RN, Aung T, Medeiros FA. The pathophysiology and treatment of glaucoma: a review. JAMA. 2014;311(18):1901–1911. doi:10.1001/jama.2014.3192

10. Bowd C, Zangwill LM, Weinreb RN, Medeiros FA, Belghith A. Estimating Optical Coherence Tomography Structural Measurement Floors to Improve Detection of Progression in Advanced Glaucoma. Am J Ophthalmol. 2017;175:37–44. doi:10.1016/j.ajo.2016.11.010

11. Musch DC, Lichter PR, Guire KE, Standardi CL. The Collaborative Initial Glaucoma Treatment Study: study design, methods, and baseline characteristics of enrolled patients. Ophthalmology. 1999;106(4):653–662. doi:10.1016/s0161-6420(99)90147-1

12. The Advanced Glaucoma Intervention Study (AGIS): 7. The relationship between control of intraocular pressure and visual field deterioration.The AGIS Investigators. Am J Ophthalmol. 2000;130(4):429–440. doi:10.1016/s0002-9394(00)00538-9

13. Castro VM, Gainer V, Wattanasin N, et al. The Mass General Brigham Biobank Portal: an i2b2-based data repository linking disparate and high-dimensional patient data to support multimodal analytics. J Am Med Inform Assoc. 2022;29(4):643–651. doi:10.1093/jamia/ocab264

14. Kerr KF, McClelland RL, Brown ER, Lumley T. Evaluating the incremental value of new biomarkers with integrated discrimination improvement. Am J Epidemiol. 2011;174(3):364–374. doi:10.1093/aje/kwr086

15. Gharahkhani P, Jorgenson E, Hysi P, et al. Genome-wide meta-analysis identifies 127 open-angle glaucoma loci with consistent effect across ancestries. Nat Commun. 2021;12(1):1258. doi:10.1038/s41467-020-20851-4

16. Mbatchou J, Barnard L, Backman J, et al. Computationally efficient whole-genome regression for quantitative and binary traits. Nat Genet. 2021;53(7):1097–1103. doi:10.1038/s41588-021-00870-7

17. Chang CC, Chow CC, Tellier LC, Vattikuti S, Purcell SM, Lee JJ. Second-generation PLINK: rising to the challenge of larger and richer datasets. Gigascience. 2015;4:7. doi:10.1186/s13742-015-0047-8

18. Garway-Heath DF, Poinoosawmy D, Fitzke FW, Hitchings RA. Mapping the visual field to the optic disc in normal tension glaucoma eyes. Ophthalmology. 2000;107(10):1809–1815. doi:10.1016/s0161-6420(00)00284-0

19. Harwerth RS, Wheat JL, Fredette MJ, Anderson DR. Linking structure and function in glaucoma. Prog Retin Eye Res. 2010;29(4):249–271. doi:10.1016/j.preteyeres.2010.02.001

20. Malik R, Swanson WH, Garway-Heath DF. “Structure-function relationship” in glaucoma: past thinking and current concepts. Clin Exp Ophthalmol. 2012;40(4):369–380. doi:10.1111/j.1442-9071.2012.02770.x

21. Hood DC, Raza AS, de Moraes CGV, Liebmann JM, Ritch R. Glaucomatous damage of the macula. Prog Retin Eye Res. 2013;32:1–21. doi:10.1016/j.preteyeres.2012.08.003

22. Wiggs JL, Hauser MA, Abdrabou W, et al. The NEIGHBOR consortium primary open-angle glaucoma genome-wide association study: rationale, study design, and clinical variables. J Glaucoma. 2013;22(7):517–525. doi:10.1097/IJG.0b013e31824d4fd8

23. Han X, Gharahkhani P, Hamel AR, et al. Large-scale multitrait genome-wide association analyses identify hundreds of glaucoma risk loci. Nat Genet. 2023;55(7):1116–1125. doi:10.1038/s41588-023-01428-5

24. Wiggs JL, Pasquale LR. Genetics of glaucoma. Hum Mol Genet. 2017;26(R1):R21-R27. doi:10.1093/hmg/ddx184

25. Weinreb RN, Leung CKS, Crowston JG, et al. Primary open-angle glaucoma. Nat Rev Dis Primers. 2016;2:16067. doi:10.1038/nrdp.2016.67

26. Gardiner SK, Fortune B, Demirel S. Signal-to-Noise Ratios for Structural and Functional Tests in Glaucoma. Transl Vis Sci Technol. 2013;2(6):3. doi:10.1167/tvst.2.6.3

27. Chen L, Zhao Y, Hashemabad SK, et al. Deep-learning-derived glaucoma-related endophenotypes enable novel genome-wide genetic and functional discovery. medRxiv. Published online November 6, 2025:2025.11.04.25339517. doi:10.1101/2025.11.04.25339517

