## Supplemental Figures for "Spatial Decomposition of Longitudinal RNFL Maps Reveals Distinct Modes of Glaucomatous Progression with Structure–Function and Genetic Signatures"

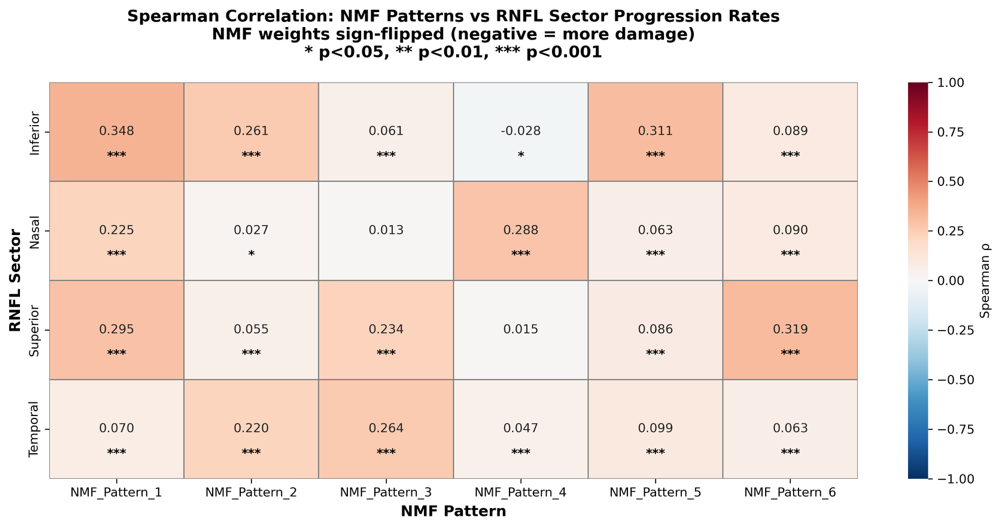

**Figure S1.** A heatmap showing Spearman rank correlations between pattern coefficients and standard sectoral RNFL thinning rates in the independent validation set. * P < 0.05, **P < 0.01, *** P < 0.001.

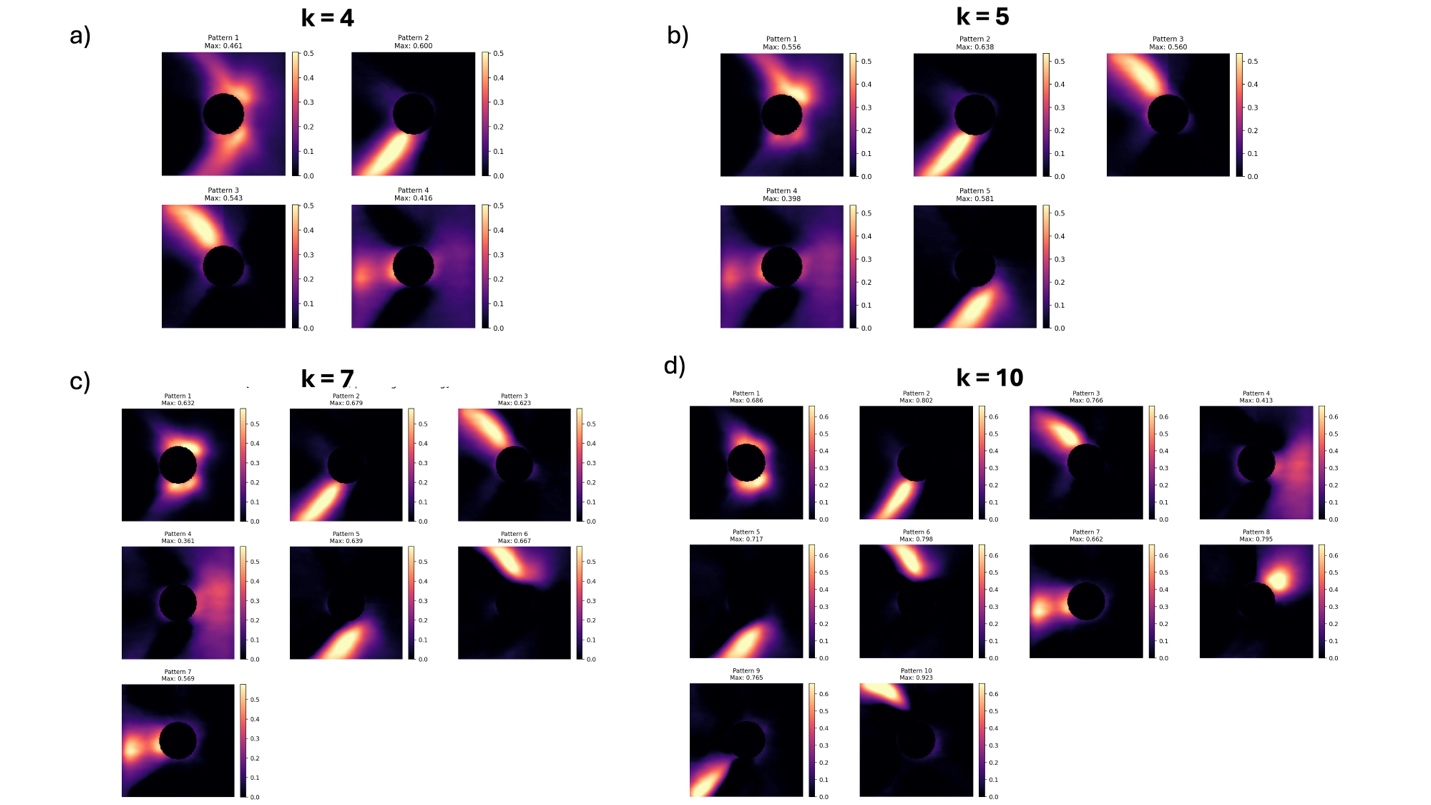

**Figure S2.** Spatial basis patterns derived from non-negative matrix factorization (NMF) of RNFL maps at (a) k = 4, (b) k = 5, (c) k = 7, and (d) k = 10. Each heatmap displays a learned spatial pattern, with brighter regions indicating areas of greater contribution to that pattern. The central dark circle represents the masked optic disc region. At k = 4, distinct inferior arcuate bundle defects are merged into a single component. At k = 5, superior and inferior hemiretinal patterns emerge but focal peripapillary and distal arcuate defects remain combined. At k = 7 and k = 10, anatomically coherent patterns begin to fragment into redundant sub-components (e.g., multiple patterns capturing overlapping regions of the inferior arcuate bundle) without improved anatomical specificity. The selected solution (k = 6, shown in Figure 1 in main text) achieved maximal separation of clinically recognizable nerve fiber bundle trajectories, distinguishing focal peripapillary from distal arcuate defects in both hemifields while avoiding over-fragmentation.

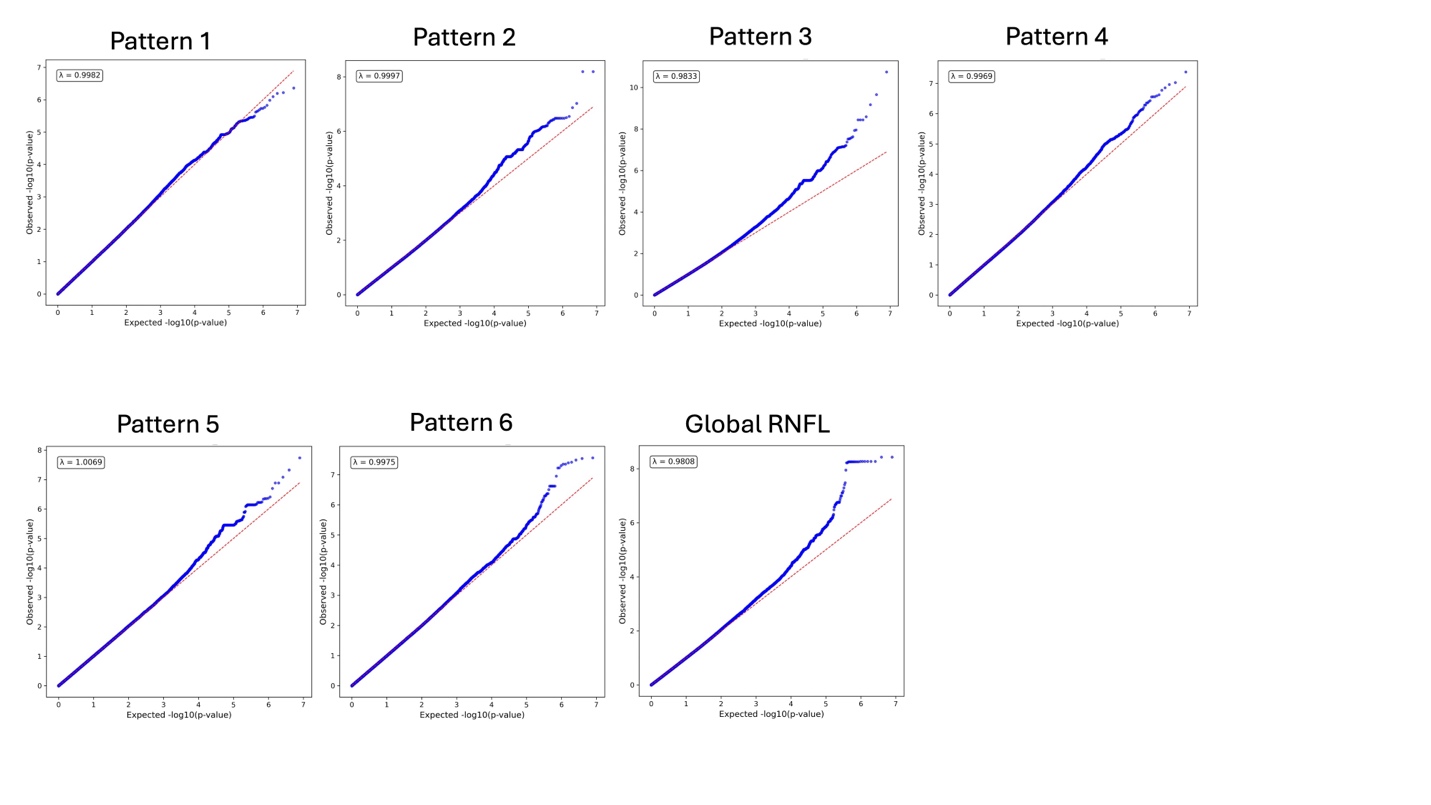

**Figure S3.** Q-Q plots comparing observed versus expected −log₁₀(P-values) under the null hypothesis of no association for each of the six NMF-derived spatial progression patterns (Patterns 1–6) and global RNFL thinning rate. The red dashed line represents the expected distribution under the null. Genomic inflation factors (λ) are displayed for each phenotype. All λ values were close to 1.0, indicating adequate control of population stratification and no systematic inflation of test statistics.
