## Supplemental Methods for "Spatial Decomposition of Longitudinal RNFL Maps Reveals Distinct Modes of Glaucomatous Progression with Structure–Function and Genetic Signatures"

**Supplementary Method**

**Registration of longitudinal RNFL thickness maps**

To enable pixel-wise analysis of retinal nerve fiber layer (RNFL) thickness changes over time, we developed a two-stage registration pipeline that aligns longitudinal optical coherence tomography (OCT) scans to a common coordinate frame for each eye.

In stage 1, we performed rigid and affine registration using the en face OCT fundus images (200 × 200 pixels) extracted from the Cirrus HD-OCT optic disc cube scans. We employed SuperRetina,^[cite] a deep learning–based retinal image feature extractor pretrained on fundus photographs, to detect keypoints and compute local descriptors for each image. For each eye, the scan with the greatest number of detected keypoints across all available timepoints was designated as the fixed (reference) frame. This approach maximizes the likelihood of successful feature matching across the longitudinal series. Correspondences between moving and fixed images were established using a two-tier matching strategy. The primary method required mutual nearest-neighbor consistency combined with Lowe's ratio test (threshold = 0.9) to ensure high-confidence matches. When insufficient matches were obtained, a fallback to ratio-test–only matching was employed. Geometric transforms were estimated using random sample consensus (RANSAC) with a hierarchical model selection approach. We sequentially tested similarity (4 degrees of freedom), affine (6 degrees of freedom), and projective (homography, 8 degrees of freedom) transforms, selecting the model that yielded the greatest number of inliers while passing geometric sanity checks. Homography transforms exhibiting excessive perspective distortion were rejected in favor of lower-order models when available. Registered images were rejected if the out-of-field-of-view area exceeded 10% of the total image area, indicating failed or unreliable registration.

The geometric transforms derived from en face image registration were scaled to match the dimensions of the corresponding RNFL thickness maps and applied using bicubic interpolation. For left eyes, transforms were conjugated with horizontal reflection matrices to account for the prior harmonization of all RNFL maps to a right-eye orientation. To facilitate cross-sectional comparisons and ensure consistent spatial correspondence of the optic nerve head across eyes, we applied an additional translation to align all eyes to a common disc center. The target center was defined as the median optic nerve head centroid position (derived from automated Cirrus segmentation) across all eyes in the cohort, computed in the harmonized (right-eye) coordinate frame. Scans with optic nerve head centers displaced more than 30 pixels from the image center were excluded to avoid edge artifacts. The registration pipeline produced spatially aligned longitudinal RNFL thickness maps for each eye, with consistent optic disc positioning across eyes, enabling pixel-wise computation of RNFL thinning rates for subsequent pattern analysis.

**Quality control of genotype data**

Samples were excluded for genotyping call rate <95%, discordant sex, or outlying heterozygosity (>3 standard deviations from the mean). Variants were excluded for imputation quality score (R²) < 0.6, call rate <98%, minor allele frequency (MAF) <1%, or deviation from Hardy-Weinberg equilibrium (P < 1 × 10-8). To account for population stratification, principal component analysis was performed on a linkage disequilibrium (LD)-pruned set of common variants (MAF > 1%, r² < 0.1, 500 kb window) using PLINK v2.0. Covariates in all genetic association analyses include sex, age, age squared, genotyping array/batch, and the first 10 genomic principal components.
